# Cerebral venous sinus thrombosis (CVST) is not significantly linked to COVID-19 vaccines or non-COVID vaccines in a large multi-state US health system

**DOI:** 10.1101/2021.04.20.21255806

**Authors:** Colin Pawlowski, John Rincón-Hekking, Samir Awasthi, Viral Pandey, Patrick Lenehan, AJ Venkatakrishnan, Sairam Bade, John C. O’Horo, Abinash Virk, Melanie D. Swift, Amy W. Williams, Gregory J. Gores, Andrew D. Badley, John Halamka, Venky Soundararajan

## Abstract

Cerebral venous sinus thrombosis (CVST) has been reported in a small number of individuals who have received the mRNA vaccines^1^ or the adenoviral vector vaccines for COVID-19 in the US^2^ and Europe^3^. Continued pharmacovigilance is integral to mitigating the risk of rare adverse events that clinical trials are underpowered to detect, however, these anecdotal reports have led to the pause or withdrawal of some vaccines in many jurisdictions and exacerbated vaccine hesitancy at a critical moment in the fight against the COVID-19 pandemic. We investigated the frequencies of CVST seen among individuals who received FDA-authorized COVID-19 vaccines from Pfizer-BioNTech (n = 94,818 doses), Moderna (n = 36,350 doses) and Johnson & Johnson - J&J (n = 1,745 doses), and among individuals receiving one of 10 FDA-approved non-COVID-19 vaccines (n = 771,805 doses). Comparing the incidence rates of CVST in 30-day time windows before and after vaccination, we found no statistically significant differences for the COVID-19 vaccines or any other vaccines studied in this population. In total, we observed 3 cases of CVST within the 30 days following Pfizer-BioNTech vaccination (2 females, 1 male; Ages (years): [79, 80, 84]), including one individual with a prior history of thrombosis and another individual with recent trauma in the past 30 days. We did not observe any cases of CVST among the patients receiving Moderna or J&J vaccines in this study population. We further found the baseline CVST incidence in the study population between 2017 and 2021 to be 45 to 98 per million patient years. Overall, this real-world evidence-based study highlights that CVST is rare and is not significantly associated with COVID-19 vaccination. In addition, there is a need for a concerted international effort to monitor EHR data across diverse patient populations and to investigate the underlying biological mechanisms leading to these rare clotting events.

## Introduction

As of April 12, 2021 more than 96 million doses of the Pfizer-BioNTech mRNA vaccine, 83 million doses of the Moderna mRNA vaccine, and 6 million doses of the Johnson & Johnson (Janssen) adenovirus vectored vaccine have been administered in the United States^4^. In six patients who received the J&J vaccine (reporting rate 0.87 per million) there have been reports of the development of a rare and dangerous adverse event, cerebral venous sinus thrombosis (CVST) associated with thrombocytopenia, in the weeks following vaccine administration^5,6^. All six cases occurred among women between the ages of 18 and 48, a high-risk group for this condition^7^, and symptoms occurred 6 to 13 days after vaccination^8^. One healthy 25-year old male who received the J&J vaccine as a part of the clinical trial also developed CVST^9^. The European Medicines Agency (EMA) is investigating an association between the AstraZeneca adenoviral vectored COVID-19 vaccine and the development of a similar thrombotic phenotype. An EMA committee carried out an in-depth review of 62 cases of cerebral venous sinus thrombosis and 24 cases of splanchnic vein thrombosis reported in the EU drug safety database (EudraVigilance) as of 22 March 2021. The cases were primarily generated via the spontaneous reporting systems of the EMA and the UK where around 25 million people have received the vaccine^10^.

Although there is no consensus regarding the causality of these associations or their appropriate impact on vaccine use^11^, this phenotype has now been dubbed vaccine-induced immune thrombotic thrombocytopenia (VITT)^12^. This name derives from the similar laboratory and clinical findings in patients with heparin induced thrombocytopenia (HIT), including widespread thrombosis in the context of declining platelets and elevated anti-platelet factor 4 (PF4) antibodies). Importantly, most individuals in whom CVST and thrombocytopenia have been reported after COVID-19 vaccination were not previously exposed to heparin, indicating that this represents a process which is pathophysiologically distinct from HIT.

Of note, concerns surrounding CVST and VITT primarily involve two adenovirus vectored vaccines and worries about a potential class effect. So far similar concerns have not been raised with regard to the two mRNA vaccines authorized for use in the United States (Pfizer/BioNTech and Moderna). However, there are reports of CVST occurring in individuals after receiving the mRNA vaccines^12^, and one group found a 4 in 1 million risk of developing CVST following receipt of an mRNA vaccine in their study cohort^13^. The accumulation of case reports has led to pauses and use restriction of adenoviral vectored vaccines in the United States and Europe, which could have major implications for controlling the COVID-19 pandemic. In light of this, it is critical to understand whether the risk of developing CVST is actually higher after receiving a COVID-19 vaccine than before, and specifically whether any COVID-19 vaccines (mRNA or adenoviral) demonstrably increase the risk of developing CVST and/or VITT. This study investigates these questions by leveraging a large multi-state EHR system to analyze individuals who have received over 130,000 doses of COVID-19 vaccines and over 771,000 doses of non-COVID-19 vaccines.

## Methods

### Institutional Review Board (IRB)

This is a retrospective study of individuals who underwent polymerase chain reaction (PCR) testing for suspected SARS-CoV-2 infection at the Mayo Clinic and hospitals affiliated with the Mayo Clinic Health System. This study was reviewed by the Mayo Clinic Institutional Review Board (IRB) and determined to be exempt from the requirement for IRB approval (45 CFR 46.104d, category 4). Subjects were excluded if they did not have a research authorization on file.

### Study design, setting and population

This is a retrospective study of individuals who were vaccinated in the Mayo Clinic hospital system between January 1, 2017 and March 15, 2021 and also received at least one SARS-CoV-2 PCR test. The outcome of interest was cerebral venous sinus thrombosis, identified either by the presence of a corresponding ICD code (I67.6, I63.6, O22.5, G08.X) or by an NLP algorithm that detected a positive diagnosis of CVST of synonyms concepts in their clinical notes (eMethods in Supplement). Clinicians manually reviewed the patients identified as positive for CVST to confirm that it was an acute diagnosis. Subjects were excluded if they did not have a research authorization on file. No subjects were excluded on the basis of demographics, comorbidities, or other clinical characteristics.

### Incidence rates

Incidence rates of first-time CVST were calculated as the number of events observed within a given time period divided by the total number of patient-years in the observation period. For the calculation of background incidence rates from 2017 to present, each of the 30-day windows following the 771,305 vaccine administrations were removed from the total number of patient-years of observation; any CVST events occurring within these windows were not counted towards the background incidence rate and the corresponding patients were removed from the cohort at the time of CVST occurrence. For the calculation of incidence rates within vaccine risk windows, the observation period was defined as the 1–30-day period following vaccine administration. Confidence intervals are *1*.*96 x standard error (SE)* where *SE = (N/t*^*2*^)^*0*.*5*^; *N* is the number of CVST events and *t* is the observation period duration multiplied by the number of patients under observation.

### Statistical analysis

We described the characteristics and frequency of cases of CVST stratified by vaccine. For each vaccine, we reported the number of patients newly diagnosed with CVST for the following time periods: +1 to +30 days after the vaccine dose, +1 to +15 days after the vaccine dose, −30 to −1 days before the vaccine dose, and −15 to −1 days before the vaccine dose. We set an at-risk window for CVST post-vaccination to 30 days as all cases reported thus far in the literature and VAERS fell within this window. We defined the relative risk to be post-vaccination incidence in the (+1 to +30 days) divided by the pre-vaccination incidence (−30 to −1 days), and we reported 95% confidence intervals for this metric. In addition, we computed Fisher exact test p-values for the null hypothesis that the relative risk is equal to one. Analyses were conducted in the Python programming language.

### Augmented curation of unstructured clinical notes

We used artificial intelligence (AI) driven augmented curation of EHR clinical notes from 601,741 patients between January 1, 2017 and April 15, 2021 from the Mayo Clinic health system to determine CVST diagnoses and other comorbidities. The augmented curation approach has been detailed previously^14^. Briefly, we used previously developed and detailed state-of-the-art BERT-based neural networks to rapidly curate clinical notes that were authored within 6 months of and COVID-19 diagnoses. Specifically, the model extracts sentences containing clinical phenotypes and symptoms and classifies their sentiment into the following categories: Yes (confirmed clinical manifestation or diagnosis), No (ruled out clinical manifestation or diagnosis), Maybe (possibility of clinical manifestation or diagnosis), and Other (alternate context, e.g., family history of disease). The neural networks are pre-trained on 3.17 billion tokens from the biomedical and computer science domains (SciBERT) and subsequently trained using 18,490 sentences and approximately 250 phenotypes with an emphasis on cardiovascular, pulmonary, and metabolic phenotypes. It achieves 93.6% overall accuracy and over 95% precision and recall for both “Yes” and “No” sentiment classification.

### Vaccine Adverse Event Reporting System review

We also looked at the data collected by Vaccine Adverse Event Reporting System (VAERS)^15^. We searched for records of any vaccine that had any of the following adverse events: (10007830) Cavernous Sinus Thrombosis, (10061251) Intracranial Venous Sinus Thrombosis, (10042567) Superior Sagittal Sinus Thrombosis, (10008138) Cerebral Venous Thrombosis, (10083037) Cerebral Venous Sinus Thrombosis, (10044457) Transverse Sinus Thrombosis within 30 days. As of April 17, 2021, VAERS has data up to April 10, 2021.

## Results

We determined the background incidence rate of CVST at the Mayo Clinic, defined as the incidence rate of CVST excluding all 30 day time windows following the administration of any non-SARS-CoV-2 vaccines (266,094 unique patients, 771,805 vaccination events). In total, 165 CVST events were identified. The incidence rate ranged from 45 to 98 per million patient years between 2017 and 2021 (Figure 1A). We then investigated the occurrence of CVST in individuals receiving a COVID-19 vaccine or various non-COVID-19 vaccines. Of the 132,916 total COVID-19 vaccine doses administered, 1,745 were the Janssen vaccine, 36,352 were the Moderna and 94,819 the Pfizer/BioNTech. 771,805 non-COVID-19 vaccine doses were captured in our cohort. We computed annual CVST incidence rates in the non-SARS-CoV-2 vaccine risk window (Figure 1B) and the incidence rate to-date in the SARS-CoV-2 (Figure 1C) vaccine risk window. These incidence rates ranged from 0 to 125 per million patient years for non-SARS-CoV-2 vaccines and 292 per million patient years for SARS-CoV-2 vaccines, though the standard errors in these cases were larger than the computed incidence rates. Among the 165 cases of CVST identified since 2017, four patients also had thrombocytopenia (platelets < 150k/ul) within 3 days of CVST diagnosis, for an incidence rate of 2 per million patient years. In three cases, platelets were in the 133k-146k/ul range, and in one case, platelets were in the 74k-84k/ul range. None of these cases occurred in the 30 days following a vaccination of any type.

**Figure 1.**
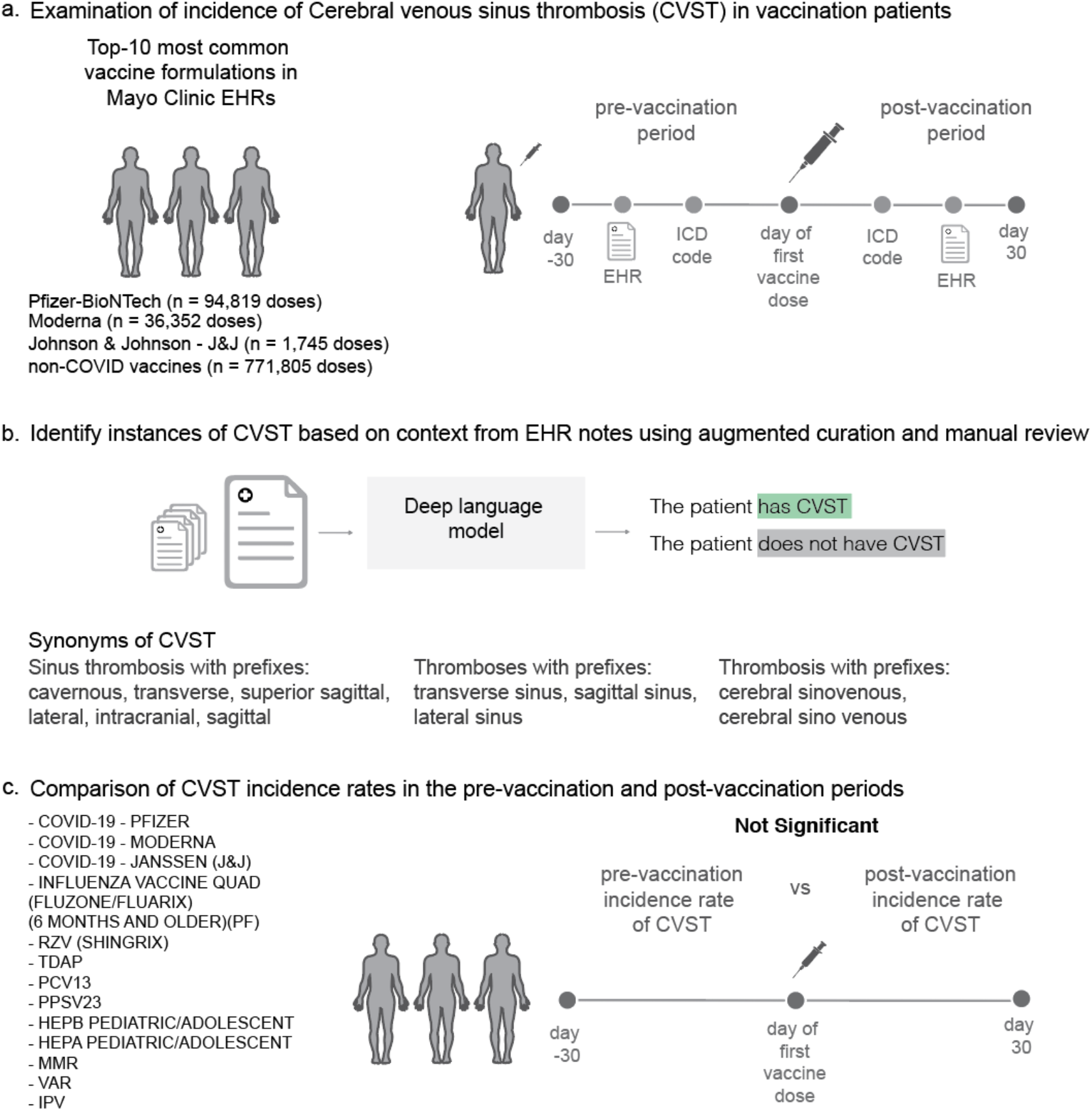
Overview of the analysis of Cerebral venous sinus thrombosis (CVST) from the EHR database of a multi-state health system. (a) Examination of incidence of Cerebral venous sinus thrombosis (CVST) in vaccination patients. (b) Identify instances of CVST based on context from EHR notes. (c) Comparison of CVST incidence rates in the pre-vaccination and post-vaccination periods.

**Figure 2.**
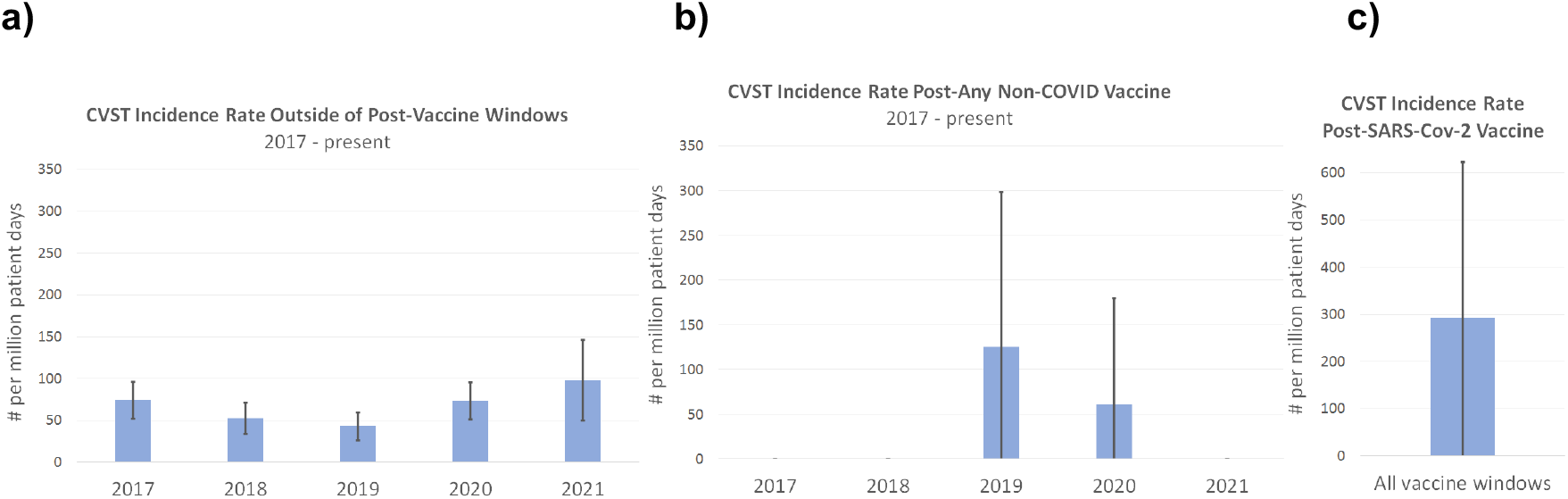
Incidence rates of cerebral venosinus thrombosis (CVST) in the study population. Individuals included in this plot are those who received a SARS-CoV-2 polymerase chain reaction (PCR) test at the Mayo Clinic in 2020-21. Incidence of CVST is defined as the first attribution of a diagnosis of CVST to a patient within a physician note, after removal of sentences referring to past occurrences of CVST. A vaccine risk window is defined as the 30 days following administration of a vaccine. **a)** The annual incidence rates of CVST outside of vaccine risk windows (top 10 non-SARS-CoV-2 vaccines and SARS-CoV-2 vaccines). **b)** The annual incidence rates of CVST in the vaccine risk window for the top 10 non-SARS-CoV-2 vaccines administered from January 1, 2017 to April 15, 2021. **c)** The incidence rate of CVST following any SARS-CoV-2 vaccine. Error bars depict standard error.

Among patients receiving any COVID-19 vaccine, there were 10 cases of CVST observed (0.0019%) in the 30 days following vaccine administration, and 10 cases of CVST were observed in the 30 days prior to administration. 3 of the 10 post-vaccination CVST cases were from individuals receiving the Pfizer/BioNTech vaccine, but there were also 3 cases among this same cohort in the 30-day pre-vaccination window, suggesting that these events were likely not caused by the vaccine (RR: 1.0, 95% CI: [0.23, 4.40] (**Table 1**). To date, no cases of CVST have been documented within 30 days before or after any doses of the Moderna or Janssen vaccines at the Mayo Clinic (**Table 1**). The relative risk of CVST in the 30 days following any COVID-19 vaccination was not statistically significant (RR: 1.50, 95% CI: [0.28, 7.10], see **Table 1**).

**Table 1:** Comparison of cerebral venous sinus thrombosis (CVST) cases recorded pre- and post-vaccination in the Mayo Clinic EHR data. Results are shown for the top-10 most common vaccine formulations in the Mayo Clinic EHR database, along with the three FDA-authorized COVID-19 vaccines (Pfizer/BioNTech, Moderna, and Janssen). Vaccine dose totals are provided for the study time period of January 1, 2017 to March 15, 2021. Neural network models applied to clinical notes were used to determine initial CVST diagnoses. For each vaccine formulation, CVST patient counts are shown for the following time periods: +1 to +30 days after the vaccine dose, −30 to −1 days before the vaccine dose, +1 to +15 days after the vaccine dose, and −1 to - 15 days before the vaccine dose. The relative risk is defined as the post-vaccination incidence (+1 to +30 days) divided by the pre-vaccination incidence (−30 to −1 days). In the last column, a p-value for Fisher exact test is shown for the null hypothesis that the relative risk is equal to one. Rows are sorted by total number of vaccine doses. Totals for all vaccines, non-COVID-19 vaccines, and COVID-19 vaccines are shown in the bottom rows in bold.

|  |  | Number of patients with initial diagnoses of CVST |  |  |  |  |  |
| --- | --- | --- | --- | --- | --- | --- | --- |
| Vaccine Name | Total Number of Doses | +1 to +30 days post-vaccination | -31 to -1 days pre-vaccination | +1 to +15 days post-vaccination | -15 to -1 days pre-vaccination | Relative Risk [95% CI] | Fisher Exact Test p-value |
| INFLUENZA VACCINE QUAD (FLUZONE/FLUARIX) (6 MONTHS AND OLDER)(PF) | 334229 | 3 | 4 | 3 | 3 | 0.75 [0.19, 3.14] | 1.00 |
| RZV (SHINGRIX) | 112062 | 0 | 2 | 0 | 1 | 0.00 [0.00, 4.17] | 0.50 |
| TDAP | 94895 | 0 | 2 | 0 | 0 | 0.00 [0.00, 4.17] | 0.50 |
| SARS-COV-2 (COVID-19) - PFIZER | 94818 | 3 | 2 | 1 | 2 | 1.50 [0.28, 7.10] | 1.00 |
| PCV13 | 78672 | 0 | 0 | 0 | 0 | NA | 1.00 |
| PPSV23 | 51746 | 0 | 1 | 0 | 0 | 0.00 [0.00, 8.18] | 1.00 |
| SARS-COV-2 (COVID-19) - MODERNA | 36350 | 0 | 0 | 0 | 0 | NA | 1.00 |
| HEPB PEDIATRIC/ADOLESCENT | 32265 | 0 | 0 | 0 | 0 | NA | 1.00 |
| HEPA PEDIATRIC/ADOLESCENT | 27390 | 0 | 0 | 0 | 0 | NA | 1.00 |
| MMR | 19560 | 0 | 0 | 0 | 0 | NA | 1.00 |
| VAR | 16368 | 0 | 0 | 0 | 0 | NA | 1.00 |
| IPV | 4618 | 0 | 0 | 0 | 0 | NA | 1.00 |
| SARS-COV-2 (COVID-19) - JANSSEN (J&J) | 1745 | 0 | 0 | 0 | 0 | NA | 1.00 |
| **All vaccines** | **904718** | **10** | **10** | **6** | **6** | **1.00 [0.43, 2.35]** | **1.00** |
| **Non-COVID-19 vaccines** | **771805** | **7** | **8** | **5** | **4** | **0.88 [0.33, 2.36]** | **1.00** |
| **COVID-19 vaccines** | **132913** | **3** | **2** | **1** | **2** | **1.50 [0.28, 7.10]** | **1.00** |

The clinical characteristics of the patients in the study population with CVST and the subset of patients with CVST following COVID-19 vaccination is provided in **Table 2**. In total, we observed 3 cases of CVST within the 30 days following Pfizer-BioNTech vaccination (2 females, 1 male; Ages (years): [79, 80, 84]), including one individual with a prior history of thrombosis and another individual with recent trauma in the past 30 days. The older ages of these patients and lack of concurrent thrombocytopenia further suggests that these events were distinct from the VITT phenotype which has been reported predominantly in younger females. As compared to all individuals who experienced CVST regardless of vaccination status, patients experiencing CVST after receiving a COVID-19 vaccine generally were older (81.4 +/-2.8 years old vs. 47.7 +/-22.3 years old) and had more comorbidities (2.3 +/-2.1 comorbidities vs. 1.3 +/-1.6 comorbidities).

**Table 2:** Clinical characteristics of CVST patients (overall and following COVID-19 vaccination). Charlson comorbidities are determined by ICD codes. Other comorbidities including autoimmune disorders, heart disease, hemoglobinopathy, hypercoagulability syndrome, inflammatory bowel disease (IBD), thrombosis, and trauma are determined from the clinical notes using deep neural network models. The phenotype for trauma is determined using clinical notes from the past 30 days, and for all other phenotypes, clinical notes or ICD codes from the past 10 years are considered (relative to the CVST diagnosis date).

| Clinical Characteristics | All CVST Patients<br>(n = 105) | CVST following<br>COVID-19<br>vaccination<br>(n = 3) |
| --- | --- | --- |
| Age in years |  |  |
| - Mean (std dev) | 52.4 (22.3) | 81.4 (2.8) |
| - < 18 | 7 (6.7%) | 0 (0.0%) |
| - 19-35 | 17 (16.2%) | 0 (0.0%) |
| - 36-49 | 20 (19.0%) | 0 (0.0%) |
| - 50-64 | 26 (24.8%) | 0 (0.0%) |
| - >= 65 | 35 (33.3%) | 3 (100.0%) |
| Sex |  |  |
| - Female | 64 (61.0%) | 2 (66.7%) |
| - Male | 41 (39.0%) | 1 (33.3%) |
| Race |  |  |
| - Asian | 4 (3.8%) | 0 (0.0%) |
| - Black / African American | 4 (3.8%) | 0 (0.0%) |
| - White / Caucasian | 92 (87.6%) | 3 (100.0%) |
| - Other | 2 (2.9%) | 0 (0.0%) |
| - Unknown | 1 (1.9%) | 0 (0.0%) |
| Ethnicity |  |  |
| - Hispanic or Latino | 5 (4.8%) | 0 (0.0%) |
| - Not Hispanic or Latino | 99 (94.3%) | 3 (100.0%) |
| - Unknown | 1 (1.0%) | 0 (0.0%) |
| BMI (kg/m<sup>2</sup>) |  |  |
| - Mean (std dev) | 28.7 (7.3) | 29.0 (9.3) |
| - Data availability | 51 (48.6%) | 3 (100.0%) |
| Comorbidity |  |  |
| - Autoimmune disorders | 7 (6.7%) | 0 (0.0%) |
| - Cancer (localized) | 6 (5.7%) | 1 (33.3%) |
| - Cancer (metastatic) | 0 (0.0%) | 0 (0.0%) |
| - Congestive heart failure | 2 (1.9%) | 0 (0.0%) |
| - Dementia | 0 (0.0%) | 0 (0.0%) |
| - Diabetes | 8 (7.6%) | 0 (0.0%) |
| - Diabetes (with complications) | 1 (1.0%) | 0 (0.0%) |
| - Heart disease | 22 (21.0%) | 2 (66.7%) |
| - Hemoglobinopathy | 0 (0.0%) | 0 (0.0%) |
| - HIV / AIDS | 0 (0.0%) | 0 (0.0%) |
| - Hypercoagulability syndrome | 10 (9.5%) | 0 (0.0%) |
| - IBD | 5 (4.8%) | 0 (0.0%) |
| - Liver disease (mild-to-moderate) | 6 (5.7%) | 1 (33.3%) |
| - Liver disease (severe) | 1 (1.0%) | 0 (0.0%) |
| - Myocardial infarction | 3 (2.9%) | 0 (0.0%) |
| - Paralysis | 0 (0.0%) | 0 (0.0%) |
| - Peptic ulcer disease | 2 (1.9%) | 0 (0.0%) |
| - Peripheral vascular disease | 7 (6.7%) | 0 (0.0%) |
| - Pulmonary disease | 10 (9.5%) | 1 (33.3%) |
| - Renal disease | 6 (5.7%) | 0 (0.0%) |
| - Rheumatic disease | 2 (1.9%) | 0 (0.0%) |
| - Stroke | 6 (5.7%) | 0 (0.0%) |
| - Thrombosis | 29 (27.6%) | 1 (33.3%) |
| - Trauma | 5 (4.8%) | 1 (33.3%) |
| Number of comorbidities |  |  |
| - Mean (std dev) | 1.3 (1.6) | 2.3 (2.1) |

## Discussion

Vaccines to prevent COVID-19 were developed and tested at an unprecedented pace over the past year. They have shown excellent effectiveness both in randomized clinical trials and in the real world setting ^16–18^. While no major safety concerns were identified during the large Phase 3 trials of each authorized vaccine, it is important that their safety is continually assessed throughout the vaccine rollout process. Recent case reports of individuals experiencing CVST with thrombocytopenia shortly after receiving an adenoviral vector COVID-19 vaccine and increased reporting of CVST in VAERS following COVID-19 vaccination as compared to historical rates (**Supplementary Table 1**) have raised safety concerns, leading to the temporary suspension of some COVID-19 vaccines awaiting causality investigations.

While it is important to consider this reporting in pharmacovigilance decision-making, there is reason to interpret such reports with caution. Spontaneous adverse event reporting is prone to reporting bias and increased event reporting with lay press attention or the introduction of a novel agent^20–22^.During this time it is critical to rapidly extract and analyze the content of EHR systems throughout the United States in order to validate or refute whether the risk for this phenotype is in fact increased by one or more authorized vaccines. In this retrospective study of vaccine recipients across a large multi-state healthcare system, we did not detect any significant association between COVID-19 vaccination status and the development of CVST.

Specifically, we found that (1) the risk of CVST was similar in the 30 days prior to COVID-19 vaccination compared to the 30 days after vaccination; (2) the risk of CVST within 30 days of COVID-19 vaccination was similar to the risk of CVST within 30 days of all analyzed non-COVID vaccinations; and (3) the risk of CVST after COVID-19 vaccination was similar to the baseline risk of CVST across a large cohort of patients in a multi-state healthcare system.

Our analysis demonstrates that to date no vaccine of any class, for COVID-19 or otherwise, has been associated with a statistically significant increased relative risk of cerebral venous sinus thrombosis in the Mayo Clinic Health System. If the average CVST incidence rate seen in our cohort (71.7 per million patient years) was extrapolated across the United States we would expect to see approximately 19 cases of CVST in the 14 days after vaccination amongst the 7 million patients who have received the Johnson & Johnson vaccine by chance alone, while there has been only 6 reported cases to date. This is in line with earlier research demonstrating that the incidence rate of venous thromboembolism was no higher than expected following viral vector vaccine administration^23^ and was similar across COVID-19 vaccines^13^. Of note, our cohort incidence rate was several times that reported in other American cohorts^24^. This is likely due to our data originating from tertiary care hospitals and our ability to find CVST diagnoses in free-text as well as recorded ICD-10 codes. Further real-world evidence studies are needed to confirm these findings, but pharmacovigilance regulators should consider such evidence when making decisions regarding the critically important vaccine roll out.

This study has several limitations. First, we analyzed data from a single health system which is demographically distinct from the broader United States population. In addition, the study population was restricted to individuals who have received at least one PCR test for SARS-CoV-2 at the Mayo Clinic, which is different from the overall vaccinated population. Second, as a retrospective study our analyses are inherently limited to only the data which was deemed necessary for collection during the clinical care of each patient. For example, platelet counts are not available for most individuals within 30 days of any vaccination and quantification of anti-PF4 antibodies are even more sparse. Other clinical covariates not included in the analysis such as oral contraceptives and smoking status may be potential confounding factors. Finally, our cohort contains 1,745 individuals who received the Johnson & Johnson (Janssen) COVID-19 vaccine. Given that VITT has only been reported in 6 of approximately 7 million recipients of this vaccine^2^, we are unlikely to detect a signal for it in this cohort.

Despite these limitations, the estimation of the baseline incidence rate for CVST, as well as for CVST with thrombocytopenia, in a cohort of over 600,000 individuals is useful to contextualize the reported frequencies of CVST and VITT in patients receiving all authorized COVID-19 vaccines. It also highlights the fact that CVST is likely an under-reported entity in the VAERS database, and that clinicians should be vigilant for and report this event after any vaccination in order to facilitate further research into the nature of vaccine associated thrombosis.

## Data Availability

After publication, the data will be made available upon reasonable requests to the corresponding author. A proposal with detailed description of study objectives and the statistical analysis plan will be needed for evaluation of the reasonability of requests. Deidentified data will be provided after approval from the corresponding author and the Mayo Clinic.

## Declaration of Interests

JCO receives personal fees from Elsevier and Bates College, and receives small grants from nference, Inc, outside the submitted work. ADB is a consultant for Abbvie, is on scientific advisory boards for nference and Zentalis, and is founder and President of Splissen therapeutics. nference collaborates with Janssen and other bio-pharmaceutical companies on data science initiatives unrelated to this study. These collaborations had no role in study design, data collection and analysis, decision to publish, or preparation of the manuscript. JRH, JCO, GJG, AWW, AV, MDS, and ADB are employees of the Mayo Clinic. This research has been reviewed by the Mayo Clinic Conflict of Interest Review Board and is being conducted in compliance with Mayo Clinic Conflict of Interest policies.

## Author Contributions

VS, CP, SA and PL conceived the study. All authors wrote sections of the manuscript and reviewed the findings. The authors from nference contributed methods, analysis, and software tools. The authors from Mayo Clinic reviewed the study design, clinical findings, and the manuscript’s health policy implications. All authors revised the manuscript based on feedback received.

## Funding statement

No external funding was received for this study.

## Supplementary Materials

**Supplementary Table 1:**
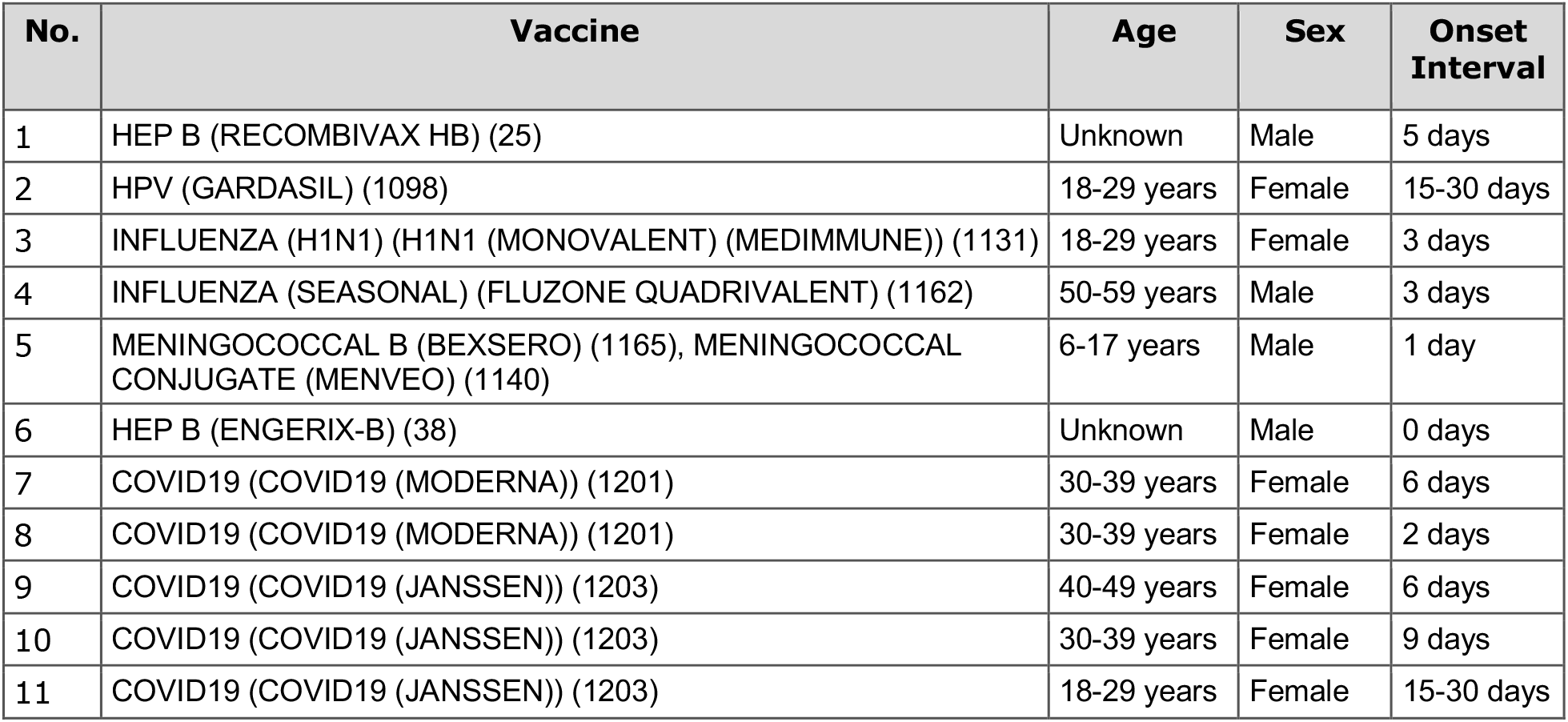
Total CVST cases that received any vaccination from VAERS Data. As of April 17, 2021, VAERS had data up to April 10, 2021. Onset Interval: 0-30 days. The following adverse events were considered: Cavernous Sinus Thrombosis, Intracranial Venous Sinus Thrombosis, Superior Sagittal Sinus Thrombosis, Cerebral Venous Thrombosis, Cerebral Venous Sinus Thrombosis, and Transverse Sinus Thrombosis.

## References

1. Vaccine-induced Immune Thrombotic Thrombocytopenia - Hematology.org. https://www.hematology.org:443/covid-19/vaccine-induced-immune-thrombotic-thrombocytopenia.

2. HAN Archive - 00442. https://emergency.cdc.gov/han/2021/han00442.asp (2021).

3. Schultz, N. H. et al. Thrombosis and Thrombocytopenia after ChAdOx1 nCoV-19 Vaccination. N. Engl. J. Med. (2021) doi:10.1056/NEJMoa2104882.

4. CDC. COVID Data Tracker. https://covid.cdc.gov/covid-data-tracker (2020).

5. Sadoff, J., Davis, K. & Douoguih, M. Thrombotic Thrombocytopenia after Ad26.COV2.S Vaccination - Response from the Manufacturer. N. Engl. J. Med. (2021) doi:10.1056/NEJMc2106075.

6. Muir, K.-L., Kallam, A., Koepsell, S. A. & Gundabolu, K. Thrombotic Thrombocytopenia after Ad26.COV2.S Vaccination. N. Engl. J. Med. (2021) doi:10.1056/NEJMc2105869.

7. Tatlisumak, T., Jood, K. & Putaala, J. Cerebral Venous Thrombosis: Epidemiology in Change. Stroke; a journal of cerebral circulation vol. 47 2169–2170 (2016).

8. Joint CDC and FDA Statement on Johnson & Johnson COVID-19 Vaccine. https://www.cdc.gov/media/releases/2021/s0413-JJ-vaccine.html (2021).

9. [No title]. https://www.cdc.gov/vaccines/acip/meetings/downloads/slides-2021-04/02-COVID-Janssen-508.pdf.

10. Pinho, A. C. AstraZeneca’s COVID-19 vaccine: EMA finds possible link to very rare cases of unusual blood clots with low blood platelets. https://www.ema.europa.eu/en/news/astrazenecas-covid-19-vaccine-ema-finds-possible-link-very-rare-cases-unusual-blood-clots-low-blood (2021).

11. Benefits of COVID-19 Vaccine AstraZeneca outweigh risks of blood clots. Reactions Weekly vol. 1848 2–2 (2021).

12. Cines, D. B. & Bussel, J. B. SARS-CoV-2 Vaccine-Induced Immune Thrombotic Thrombocytopenia. N. Engl. J. Med. (2021) doi:10.1056/NEJMe2106315.

13. Taquet, M. COVID-19 and cerebral venous thrombosis: a retrospective cohort study of 513,284 confirmed COVID-19 cases. (2021) doi:10.17605/OSF.IO/H2MT7.

14. Wagner, T. et al. Augmented curation of clinical notes from a massive EHR system reveals symptoms of impending COVID-19 diagnosis. Elife 9, (2020).

15. VAERS - Data. https://vaers.hhs.gov/data.html.

16. Oliver, S. E. et al. The Advisory Committee on Immunization Practices’ Interim Recommendation for Use of Janssen COVID-19 Vaccine — United States, February 2021. MMWR. Morbidity and Mortality Weekly Report vol. 70 329–332 (2021).

17. Baden, L. R. et al. Efficacy and Safety of the mRNA-1273 SARS-CoV-2 Vaccine. N. Engl. J. Med. 384, 403–416 (2021).

18. Polack, F. P. et al. Safety and Efficacy of the BNT162b2 mRNA Covid-19 Vaccine. N. Engl. J. Med. 383, 2603–2615 (2020).

19. Greinacher, A. et al. Thrombotic Thrombocytopenia after ChAdOx1 nCov-19 Vaccination. N. Engl. J. Med. (2021) doi:10.1056/NEJMoa2104840.

20. McConeghy, K. W., Bress, A., Qato, D. M., Wing, C. & Nutescu, E. A. Evaluation of dabigatran bleeding adverse reaction reports in the FDA adverse event reporting system during the first year of approval. Pharmacotherapy 34, 561–569 (2014).

21. McAdams, M. A., Governale, L. A., Swartz, L., Hammad, T. A. & Dal Pan, G. J. Identifying patterns of adverse event reporting for four members of the angiotensin II receptor blockers class of drugs: revisiting the Weber effect. Pharmacoepidemiology and Drug Safety vol. 17 882–889 (2008).

22. Bohn, J. et al. Patterns in spontaneous adverse event reporting among branded and generic antiepileptic drugs. Clin. Pharmacol. Ther. 97, 508–517 (2015).

23. Østergaard, S. D., Schmidt, M., Horváth-Puhó, E., Thomsen, R. W. & Sørensen, H. T. Thromboembolism and the Oxford-AstraZeneca COVID-19 vaccine: side-effect or coincidence? Lancet (2021) doi:10.1016/S0140-6736(21)00762-5.

24. Otite, F. O. et al. Trends in incidence and epidemiologic characteristics of cerebral venous thrombosis in the United States. Neurology 95, e2200–e2213 (2020).

